# Maternal diabetes and offspring use of dental services – Northern Ireland national cohort study

**DOI:** 10.1101/2022.11.24.22282719

**Authors:** Anas Salami, Ikhlas El Karim, Fionnuala Lundy, Tom Loney, Michael Donaldson, Ciaran O’Neill

## Abstract

**Introduction:** A small number of literature has posited a link between prenatal exposure to gestational diabetes mellitus and an increased risk of developmental defects in the enamel of offspring. However, the evidence remains inconclusive.

**Aims:** This study examined the relationship between the diabetes status of mothers and the use of dental services by offspring to that pregnancy.

**Material and methods:** Anonymised data from a cohort of mothers who carried a child to term in Northern Ireland between 2012 and 2017 and service use by the child were taken from administrative databases from March 2015 to September 2021. Descriptive statistics, differences in means and regression analyses were used to examine the relationship between service use and maternal diabetes status, controlling for covariates.

**Results:** In multivariate analyses that controlled inter alia for age and deprivation, diabetes status was negatively related to restoration, extraction, prevention, and total service use. In the analysis of the COVID period, pre-COVID prevention was negatively related to extractions, restorations, prevention, and services in general.

**Conclusion:** The relationship between maternal diabetic status and aspects of offspring use of dental services was contrary to that suggested in previous studies and warrants more detailed investigation using this valuable data resource.

## Introduction

Odontogenesis begins from the sixth week of foetal life and continues through gestation and after birth^1^. As a part of odontogenesis, amelogenesis can be altered by local and systemic factors^2^. Systemic factors include changes to and reduction of tissue oxygenation, exposure to gamma rays, fever, infections, vitamin A & D deficiency and gestational diabetes ^3^. These factors can influence odontogenesis and cause enamel hypoplasia or other amelogenesis defects ^2^. Gestational diabetes mellitus (GDM) - defined as carbohydrate intolerance of variable severity with onset or first recognition during pregnancy^4^ - rates have increased in recent years^5–9^. Several studies have suggested that it increases the risk of childhood diseases ^10–13^ including the tooth development process, by affecting tooth eruption and mineralisation ^14–16^. However, the effects of maternal diabetes on tooth development and the associated underlying mechanisms have not been thoroughly investigated ^17^. Epidemiologic and animal model studies have shown that hyperglycaemia changes may negatively affect oral health and aesthetics and cause tooth sensitivity and malocclusion ^18,19^. They have also been identified as risk factors for carious lesions and erosion in children’s teeth ^20^.

While caries is a largely preventable disease, it is one of the most common diseases of childhood ^21^. It affects 23% of children aged five years in England, and there exists evidence of steep socioeconomic inequalities in prevalence.^22^ The few available studies that have examined the relationship between glycaemic control and caries, focus on diabetes mellitus, have small sample sizes, have exhibited heterogeneous results and are consequently inconclusive with respect to the role of diabetes^23–25^. We hypothesise that gestational dysglycaemia – whether from GDM or pre-existing diabetes - may affect enamel formation and, consequently, the subsequent risk of dental caries and use of dental services for restorative care ^26,27^. This study aimed to examine the relationship between the diabetes status of mothers during pregnancy and the use of dental services by offspring to that pregnancy.

## Material and Methods

### Study Design and Data

The design of this longitudinal cohort study followed the guidelines published by “Strengthening the Reporting of Observational Studies in Epidemiology (STROBE) Statement”, 2007^28^. This study uses anonymised data secured from three sources: the Northern Ireland Maternity System (NIMATS), the Enhanced Prescribing Database (EPD), and the Dental Payment System (DPS). NIMATS is the regional administrative data repository for demographic and clinical information collected on mothers and their infants during antenatal check-ups, labour and delivery, and the postnatal period in Northern Ireland. The main source of data for NIMATS is the Patient Administration System which captures details of hospital service use by patients^29^. The data provides full coverage of births in Northern Ireland with the mother’s and child’s unique health and care numbers allowing linkage of mother to offspring from a pregnancy. Data from NIMATS used in this study included the outcome of antenatal tests, such as maternal GDM status, measures, and outcomes such as birth weight and vital status. Data extracted covered pregnancies from 2012 to 2017. These data were linked to prescription data for mothers held in the EPD^30^.

The EPD, uniquely in the UK, captures details of prescriptions filled in the community for individuals identifiable by their unique health and care number. The EPD takes data from the NHS Business Services Organization (BS0), which operates an electronic system that allows prescriptions successfully scanned on presentation to the dispensing pharmacist to be linked to the patient’s health and care number. It is estimated that approximately 85–90 % of all prescriptions scanned at the BSO result in usable data^31^. Prescriptions in the database are coded using British National Formulary (BNF; a standard drug reference text in the UK) codes that allow drug types to be identified in broad classes as well as by specific types such as Metformin or Insulin. In this analysis, we were interested only in those drugs for which prescriptions were filled during the pregnancy.

EPD and NIMATS data were, in turn, linked to those related to the use of publicly funded dental services in the community by children born to mothers captured in NIMATS using the child’s unique health and care number held in the DPS. In Northern Ireland, General Dental Practitioners (GDPs) are reimbursed in part on a fee-for-service basis^32^ that provides specific payments for specific activities, including restorations, preventions, examinations and extractions of deciduous teeth. GDPs submitted payment claims on a monthly basis to the business services organisation, which manages claims on behalf of the regional health service commissioner. The system categorises specific services provided to the patient by type of treatment. All claimable treatments are represented in the statement of dental remuneration by a code and each code has an associated fee. All payment claims detail the tooth/teeth treated along with the treatment code.^30^. The latter sets out terms and conditions under which payments for specific services are made and the fee that service attracts from the health service. Care used by children was examined from March 2015 to September 2021 – children born in 2012 likely only using services from 2015 onward.

Data were linked through the Northern Ireland Honest Broker Service (HBS). The HBS is the Trusted Research Environment for Health and Social Care (HSC) in Northern Ireland and is hosted within the HSC Regional Business Services Organisation (RBSO/BSO). It provides anonymised individual-level data for the purposes of research, with access only permitted in a controlled fashion via a safe research environment^30^. In addition to the information identified in NIMATS, EPD and DPS, the HBS can link a patient’s health and care number to their address and use the associated postcode to ascribe an area-based measure of deprivation using the Northern Ireland Multiple Deprivation Index^33^. To help preserve anonymity, areas are ranked no lower than by decile in terms of relative deprivation.

### Methods

Statement of Dental Remuneration (SDR) codes were used to group treatments into restorations, extractions, preventive care, and examinations. Details of the codes used for this are detailed in Appendix 1. Using EPD data, BNF codes were used to identify mothers who had filled a prescription for using Metformin or Insulin during their pregnancy. Mothers’ GDM or diabetes status, age, socioeconomic status, and their child’s age measured in September 2021 were taken from NIMATS. Full details of the specification of all variables are provided in Appendix 2. Descriptive statistics (mean, standard error and 95% confidence interval), t-tests for differences in means and regression analyses were used to examine the relationship between service use and maternal diabetes status, controlling for covariates. In multivariable analyses that examined service use, Zero-Inflated Poisson (ZIP) regression models were used to explore the relationship between service use and maternal diabetes status. The ZIP model takes account of the count nature of dental service use and over-dispersion in the data, i.e., the existence of large numbers of non-users and small numbers of heavy users. Persistent non-service users – defined as individuals who had made no contact with the service before March 2021-were used as the inflation factor in the ZIP models. Diabetes status was defined as present if the mother was diagnosed within NIMATS as having GDM or if EPD data identified her as filling a prescription for Metformin or Insulin during the pregnancy.

Northern Ireland is split into 890 spatial areas known as Super Output Areas (SOAs), with an average population of around 2,100. The Index of Multiple Deprivation (IMD) is the official measure of relative deprivation for small areas, ranking each area from the most deprived to the least deprived based on the rates of income, employment, health and disability, education, skills and training, barriers to housing and services, living environment and crime. ^34^ This study uses the latest IMD released from the Northern Ireland Index of Multiple Deprivation (NIMDM) 2017.^35^ The ranked scores of the IMD were transformed into IMD quintiles, ranging from the 20% of people living in the most deprived areas to the 20% least deprived areas.

The COVID pandemic resulted in a significant disruption to dental services in Northern Ireland in 2020 and subsequent to that. This is clearly seen in (Fig 1) by the sharp fall in claims from week 12 (week ending March 20th) onward in 2020 compared to 2019 and the subsequent slow recovery in claims in 2021. To accommodate the disruption to services arising from the COVID-19 pandemic, relationships were examined across time periods differentiated by the “lockdown,” i.e., the introduction of stay-at-home orders - as it applied to Northern Ireland on March 24th. The pre-Lockdown (Pre-LD) period was from January 2015 – March 2020, a total of 63 months, and the post-lockdown (Post-LD) was from April 2020 – September 2021, a total of 27 months. The actual lockdown period was excluded from the analysis. To explore relationships further, we examined the impact of maternal diabetes status controlling for pre-lockdown consumption of prevention on post-lockdown consumption of treatment services. Consumption of services was measured as the cumulative total monthly claims, and pre-lockdown prevention was calculated as the number of preventive items claimed for a child. To help sharpen focus, we restricted comparison to the oldest group of children in our study – those born in 2012 – for whom follow-up was longest and excluded those who had made no contact with the service prior to March 2020, who might be seen as habitual non-users and different in nature to other patients. Given the data distribution – a large number of zeros with a heavy tail - a generalised linear regression model was used to examine the relationship between diabetes status and the value of monthly claims by controlling for previous consumption of preventive services. A Poisson family and log link were used in the function informed by the outcome of a modified Park test and Link test^33^. All statistical analyses were performed using STATA SE 17.0 (Stata Corp). The P values reported are two-tailed, and statistical significance was set at α=0.05. Data were collected from HBS data resources; thus, no ethical approval was necessary. This study was exempted from ethical approval by Queen’s University-Belfast, United Kingdom, with reference number (QUB Ref: RGE 22_07).

**Figure 1.**
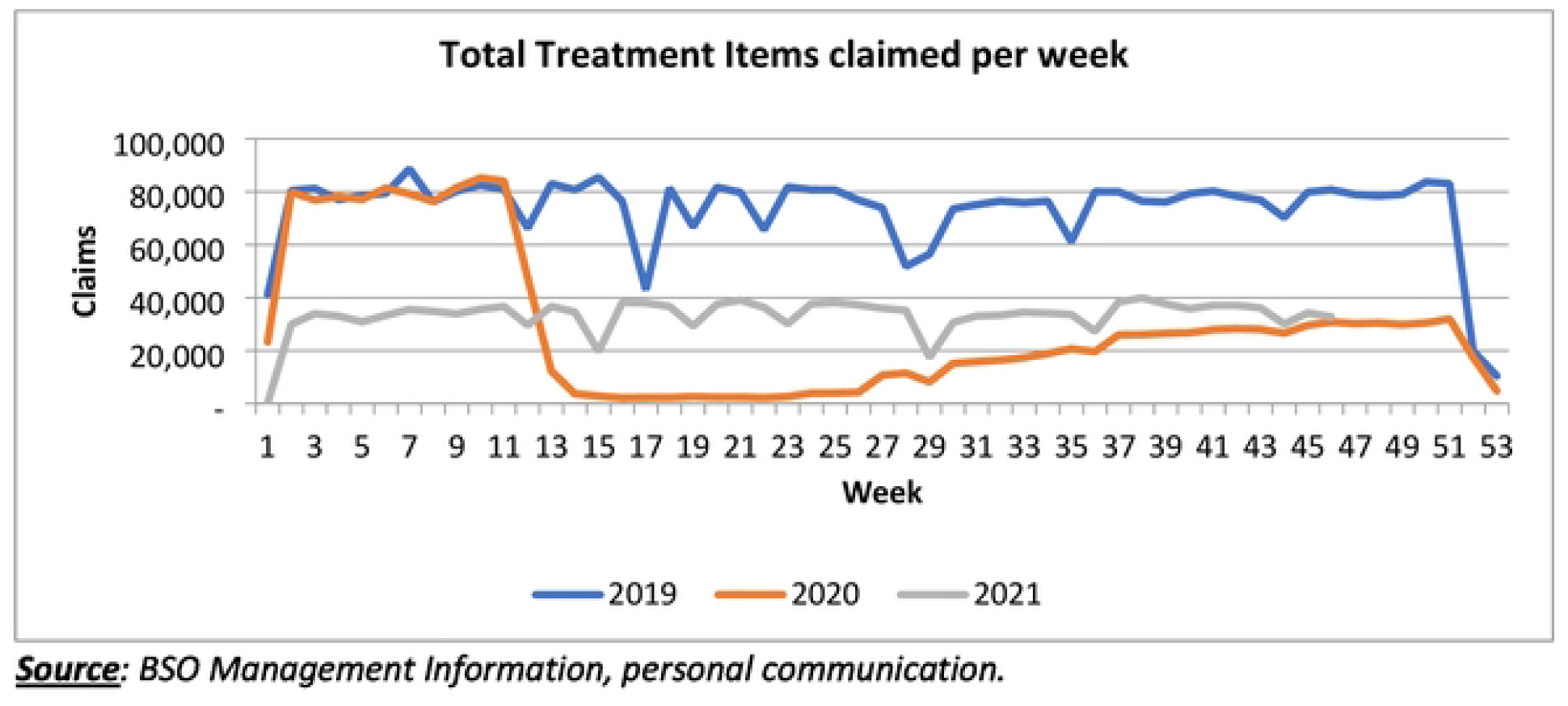
Service use before and after the COVID-19 pandemic.

## Results

A total of 144,599 mother-child dyads were included in this study. Out of this, 9,138 (6.31%) of the mothers were diagnosed as having GDM or having filled a prescription for Metformin or Insulin during their pregnancy. We also found that (80%) of children had not used dental services with respect to primary teeth by September 2021. Descriptive statistics for the sample (to two decimal places) are presented in Table 1.

**Table 1.** Descriptive Statistics.

| **N=144,599** | **Mean (±SD)** | **%** |
| --- | --- | --- |
| **Diabetes** | 0.0631 (±0.2433) | 6.31 |
| **Pre-Lockdown non-user** | 0.8001 (±0.3998) | 80 |
| **Child's age (month)** | 81.77 (±7.48) | NA |
| **DEPRIVATION Q 1** | 0.2273 (±0.4190) | 22.7 |
| **DEPRIVATION Q 2** | 0.2171 (±0.4123) | 21.71 |
| **DEPRIVATION Q 3** | 0.2065 (±0.4048) | 20.65 |
| **DEPRIVATION Q 4** | 0.1939 (±0.3953) | 19.39 |
| **DEPRIVATION Q 5** | 0.1550 (±0.3619) | 15.5 |

Table 2 reports mean differences in the number of claims between the two groups of child-mother dyads (Diabetic – Non-Diabetic) in relation to the consumption of different dental services, separating results across the pre- and post-pandemic periods. With respect to examinations, for example, the mean number of claims among children born to diabetic mothers was 0.083 in the pre-lockdown period compared to 0.089 among children born to non-diabetic mothers.

**Table 2.**
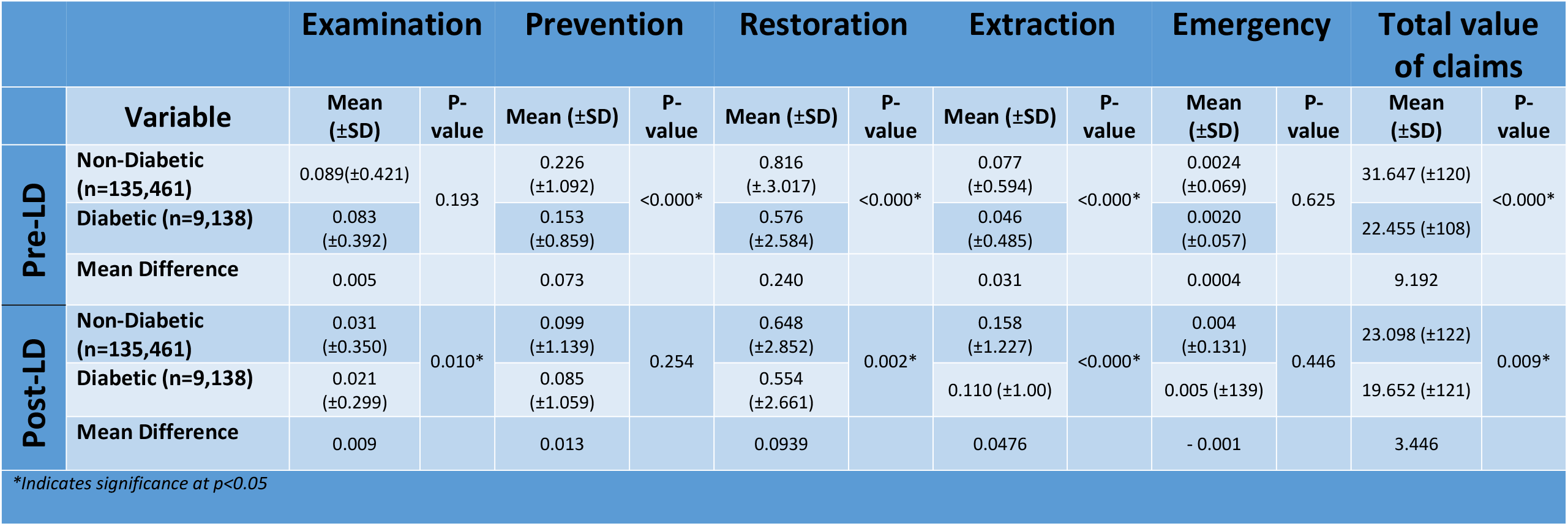
The difference in service use between diabetic and non-diabetic groups in the pre-and post-Lockdown period.

As seen from Table 2, children born to diabetic mothers showed no differences regarding the number of claims submitted for examinations or emergency visits pre-LD. On average, children of mothers with diabetes were the subject of significantly (p-value <0.000) lower claims for prevention, restorations, extraction, and the value of all care (Total value of claims) with means differences of 0.073, 0.240, 0.031, and £9.19, respectively. Differences concerning the post-lockdown period are shown through the disruption to services that must be born in mind when interpreting results. During the post-LD period, differences in prevention, restoration, extraction, and total use of services were statistically significant between the two groups, with a p-value <0.001, where children born to mothers with diabetes had fewer associated claims for services. Post-LD, the same relationships were found with restoration and extraction though no statistically significant difference was detected with prevention. Differences in emergency treatment use were consistently not significant.

The impact of the pandemic on the use of specific services across children of diabetic and non-diabetic mothers can be seen in Table 3. For example, allowing for the different duration of the two periods (63 months pre-lockdown and 27 months post-lockdown), it can be seen that among children of non-diabetic mothers, claims for examinations per month fell by approximately 20% ((0.089/63) = 0.01413 (0.031/27) = 0.001148; 0.001148/0.001413 = 0.81), while claims for extractions increased by almost 480%. Claims for emergencies and restoration also increased markedly, while those for prevention remained largely unchanged. Among children of diabetic mothers, claims for examinations post-lockdown were roughly 60% per month what they had been before lockdown. In comparison, those for extractions rose by almost 560% and emergency visits by over 580%.

**Table 3.**
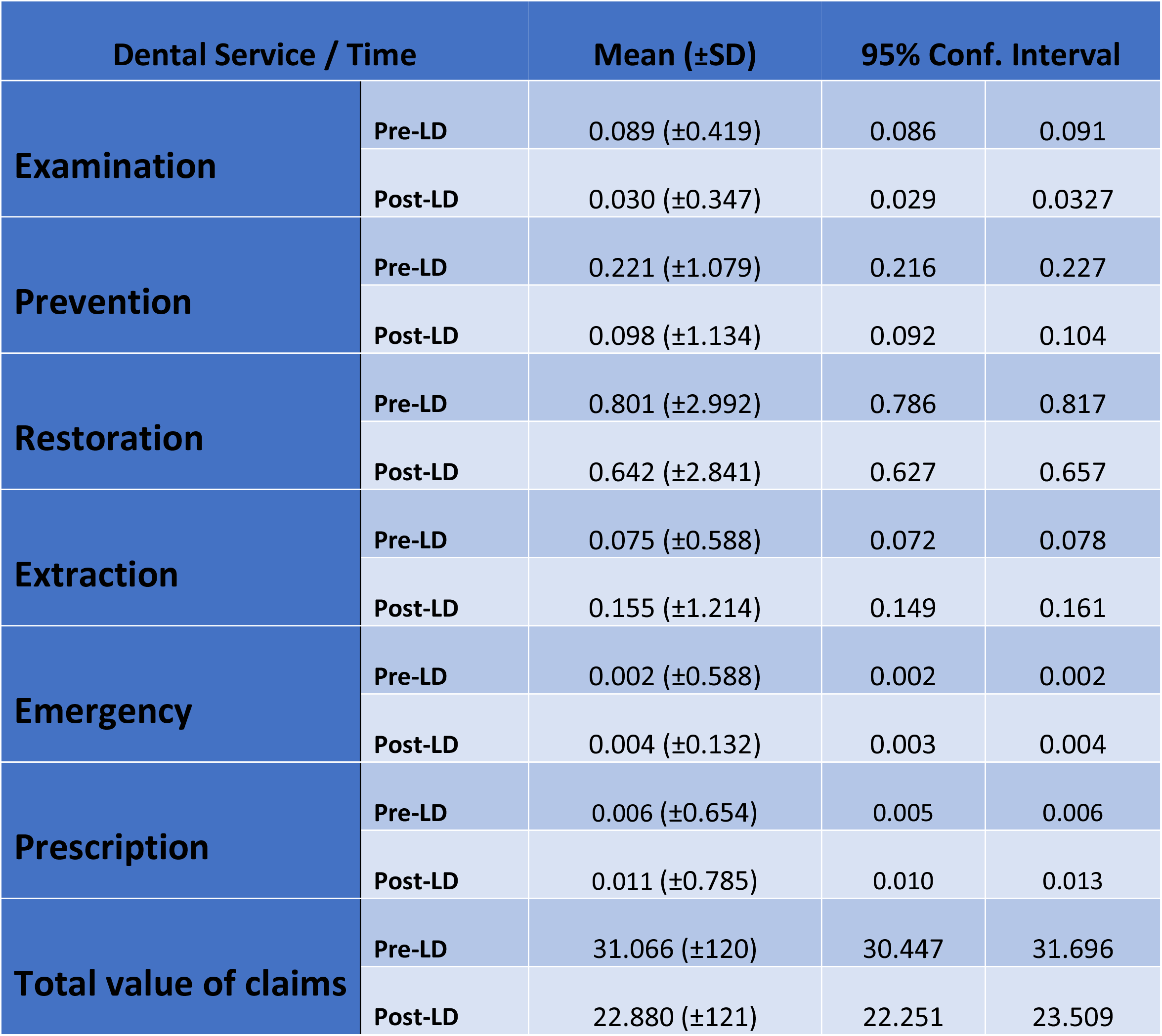
Mean difference in total service use pre- and post-lockdown.

The results of the ZIP models for claims for examinations, prevention, restoration, and extractions are presented in Table 4. When we examined the effect of the use of prevention services before the pandemic and the subsequent use of dental services after the LD period, we found that pre-LD prevention was negatively related to prevention (−0.1822), restoration (−0.0539) and extraction (−0.0358) and were statistically significant. Diabetes, in this context, did not play a significant role (Table 4). The deprivation quintile showed disparate findings across different services post-LD. As shown, for examinations, a socioeconomic gradient was evident, with those in a higher socioeconomic group (based on quintile) generating a greater number of claims for examinations than those in a lower socioeconomic group. Concerning prevention, more claims were evident among those from higher socioeconomic groups relative to the lowest. However, the difference was not as marked as for examinations compared to prevention, and the gradient was less evident among those in higher socioeconomic groups. For restorations, those in the highest socioeconomic group generated significantly fewer claims post-lockdown, controlling for other variables, than those in the lowest socioeconomic group. A similar result is evident concerning extractions where those in deprivation quintiles 2, 4, and 5 were significantly lower than quintile 1.

**Table 4.** Effect of pre-LD prevention on service use during post-LD (ZIP Analysis)

| **Impact of Pre-LD Prevention on Post-LD Dental Services Use** |  |  |  |  |
| --- | --- | --- | --- | --- |
| **N=144,599** | **Examination** | **Prevention** | **Restoration** | **Extraction** |
| **Variable** | **Coef.** | **Coef.** | **Coef.** | **Coef.** |
| **Child Age** | 0.0167* | 0.0255* | 0.0051* | 0.0168* |
| **Pre-LD Prevention** | 0.0548 | - 0.1822* | - 0.0539* | - 0.0358* |
| **DEPRIVATION Q 2** | 0.2789* | 0.1145* | 0.0115 | - 0.0666* |
| **DEPRIVATION Q 3** | 0.3252* | 0.1473* | 0.0162 | - 0.0303 |
| **DEPRIVATION Q 4** | 0.3577* | 0.1377* | - 0.0221 | - 0.0425 |
| **DEPRIVATION Q 5** | 0.3738* | 0.1444* | - 0.0914* | - 0.1620* |
| **Diabetes** | - 0.2109 | 0.0323 | - 0.0248 | - 0.0517 |
| **Pre-LD non-user** | 0.8341* | 1.1751* | 1.5735* | 1.6519* |
| *\*Indicates significance at p<0.05* |  |  |  |  |

Descriptive statistics for each model are presented in Appendix 1.

**Table 5.** Impact of Pre-LD Prevention on the total value of claims.

| **Impact of Pre-LD Prevention on the total value of claims** |  |
| --- | --- |
| **N=144,599** | **Total value of claims** |
| **Variable** | **Coef.** |
| **Child Age** | 0.0413 * |
| **Pre-LD Prevention** | -0.0393 * |
| **DEPRIVATION Q 2** | 0.0117 |
| **DEPRIVATION Q 3** | 0.0125 |
| **DEPRIVATION Q 4** | -0.1462 * |
| **DEPRIVATION Q 5** | -0.3654 * |
| **Diabetes** | 0.1082 |

*\*Indicates significance at $p < 0.05$*

When we examined the total value of claims post-LD restricted to children born in 2012 (n=11,116) and controlled by mother’s age, pre-LD prevention, deprivation quintile, and diabetes, child’s age and use of preventive services, results show that pre-LD prevention had a negative effect on the total value of claims post-LD. Older children had a lower value of claims, and higher deprivation quintiles had a higher total claim value; however, this was not statistically significant. Maternal diabetes status was negatively related to the value of claims but was not statistically significant, as presented in Table 6.

**Table 6.** Gen. Linear Model of total post-LD Total value of claims controlled by the child’s age, Deprivation Quintile, and Diabetes status of children born in 2012.

| Post-LD Total value of claims | Coef. | P>\|z | [95% Conf. Interval] |  |
| --- | --- | --- | --- | --- |
| n= 11,116 |  |  |  |  |
| Age (month) | -2.618 | 0.002* | -4.274 | -0.963 |
| Pre-LD Prevention | -10.659 | <0.000* | -14.269 | -7.050 |
| DEPRIVATION Q 2 | 2.504 | 0.787 | -15.634 | 20.642 |
| DEPRIVATION Q 3 | 9.763 | 0.280 | -7.957 | 27.4843 |
| DEPRIVATION Q 4 | 15.331 | 0.092 | -2.526 | 33.1893 |
| DEPRIVATION Q 5 | -5.154 | 0.638 | -26.647 | 16.3386 |
| Diabetes | -10.591 | 0.531 | -43.760 | 22.5777 |

## Discussion

Normal dentition starts to develop in utero, and the developing tooth bud has been shown to be sensitive to a wide range of systemic disturbances, with enamel in particular, generally unable to recover once it is damaged^36^. Enamel defects can be caused by numerous factors, including host traits, genetic factors, immunological responses to cariogenic bacteria, saliva composition, environmental and behavioural factors, and systemic diseases^37^. Several studies have suggested hyperglycaemia during pregnancy may impact enamel formation and contribute to an increased risk of caries in childhood though these were small and their findings inconclusive^1,18,19^. We studied a representative population of 144,599 mother-child dyads captured in the maternity information system in Northern Ireland between 2012 and 2017. Among mothers, 6.31% were either treated for diabetes or had a diagnosis of diabetes during the course of their pregnancy. Children of mothers with positive diabetic status consumed significantly less prevention, restoration, and extraction services compared to children of non-diabetic mothers prior to the disruption of publicly funded dental services caused by the COVID pandemic. The total value of claims for dental care submitted on behalf of children born to mothers with positive diabetic status was significantly lower during both the period that preceded the COVID lockdown and followed the COVID lockdown. At face value, this would appear to support the argument that a positive maternal diabetic status does not adversely affect offspring dental development in a manner that necessitates subsequent additional dental care. While it may or may not affect enamel development, it does not materially affect treatment needs. Some caution, however, is warranted with this interpretation. For example, while we observe the use of dental services delivered in the community by general dental practitioners, we do not observe those delivered by the Community Dental Service (CDS) or in a hospital setting. Children with a range of additional needs (which may be positively correlated with maternal diabetes status) may be more likely to be treated at the CDS. Similarly, treatments delivered in a hospital setting, such as extraction under general anaesthetic that may be related to maternal diabetes status, are not represented in the data used here.

The study, by chance, allowed us to offer some intriguing insights into the value of prevention among the children studied and into the disruption caused by the COVID pandemic. In a recent publication highlighting the indirect consequences of COVID-19, child oral health was conspicuously missing.^38^ The recent pandemic and resulting lockdowns disrupted the provision of care across various services, including community dental care in the UK. While studies have demonstrated its effectiveness in terms of activity, the impact of specific treatments or the characteristics of the affected patients has not yet been reported, specifically in Northern Ireland. As reported in our study, COVID-19 has led to an overall reduction in dental services consumption by children that have not yet recovered to pre-LD period levels. Reduction of services, especially routine examination and prevention will negatively affect children’s oral health and treatment needs in the future. This perhaps explains the significant increase by almost double in the number of claims per month in our population related to extractions, emergencies, and prescriptions post-LD. The increase in extractions could be due to worsening in the caries condition during the period of lockdown when access to services was effectively cut-off and/or individuals were hesitant to visit dental practices as it was widely publicised as a high-risk environment, which therefore increased the risk of the tooth being un-restorable. This may have longer-term effects not only in terms of oral health and occlusion but in terms of dental anxiety and behaviour.

In addition, associated with the pandemic, the number of prescriptions were significantly increased post-LD. Similar findings were reported in Scotland by Ducan et al.^39^, where they reported that antibiotic prescribing rose by 49% following the suspension of routine dental care to a peak of 34,993 prescriptions (July 2020). Their data also show that since the remobilisation of NHS dental care, antibiotic prescribing remains around 28% higher than pre-pandemic. Unnecessary and frequent prescription of antibiotics in children can lead to the development of antibiotic resistance. Antimicrobial resistance is a growing and significant global threat that has wide-ranging impacts on health, finances, food sustainability, security, environmental wellbeing, and socioeconomic development^40^. Antibiotic use in human medicine plays a crucial role in the development and spread of antimicrobial resistance^41^, and the optimisation of antibiotic use is a primary focus of the UK’s 2019–2024 national action plan to tackle antimicrobial resistance^40^.

The results of the regression analyses suggest that prevention had a valuable effect in terms of reducing the likelihood of subsequent extraction or restorative activity. As seen in Table 4, pre-COVID consumption of prevention reduced the number of claims for extraction and restorations provided post-lockdown. This effect was not seen with respect to examinations, suggesting it was not simply an artefact of the data. While the number of post-lockdown claims for prevention was also negatively related to the number provided pre-lockdown, this is to be expected given that the need for prevention where it had already been provided should be less.

While we examined a large number of mothers and children cohort in NI, which is a study strength, the limitations of our research are worth noting. For example, while we examine service use, we do not examine the treatment of specific teeth that may be particularly susceptible to maternal diabetes status. Similarly, while we control for aspects of socioeconomic status known to relate to service use, our measures are relatively crude, being area-based rather than individual-based and thus subject to potential ecological fallacy. This was a limitation imposed on us by the data; however, mindful of the potential for disclosure. Moreover, this dataset does not contain information on dental care needs. The need for dental treatment would allow for a fuller characterisation of demand and the exploration of, for example, the role of supplier-induced demand in dental treatment provision.

## Conclusion

Previous studies examining the relationship between maternal diabetic status and offspring dental caries level have produced heterogeneous results. Our study provides tentative support for the hypothesis that treatment need is not elevated as a result of maternal diabetes status though further work is needed. Our results also underscore the value of prevention, especially within a context where access to services is disrupted.

## Data Availability

All relevant data are within the manuscript and its Supporting information files.

## Acknowledgement

Authors would like to thank and acknowledge Honest Broker Services (HBS) in Northern Ireland, UK, for their continued support and for providing us with the data.

